# Genetically-proxied impaired GIPR signalling and risk of 6 cancers

**DOI:** 10.1101/2022.08.06.22278459

**Authors:** Miranda Rogers, Dipender Gill, Emma Ahlqvist, Tim Robinson, Daniela Mariosa, Mattias Johansson, Ricardo Cortez Cardoso Penha, Laure Dossus, Marc J Gunter, Victor Moreno, George Davey Smith, Richard M Martin, James Yarmolinsky

**Affiliations:** MRC Integrative Epidemiology Unit, University of Bristol, Bristol, UK; Population Health Sciences, Bristol Medical School, University of Bristol, Bristol, UK; Department of Epidemiology and Biostatistics, School of Public Health, Imperial College London, London, UK; Chief Scientific Office, Research and Early Development, Novo Nordisk, Copenhagen, Denmark; Department of Clinical Sciences, Malmö, Lund University, Malmö, Sweden; Genomic Epidemiology Branch, International Agency for Research on Cancer (IARC/WHO), Lyon, France; Nutrition and Metabolism Branch, International Agency for Research on Cancer (IARC/WHO), Lyon, France; Biomarkers and Susceptibility Unit, Oncology Data Analytics Program, Catalan Institute of Oncology (ICO), L’Hospitalet de Llobregat, Barcelona, Spain; Colorectal Cancer Group, ONCOBELL Program, Bellvitge Biomedical Research Institute (IDIBELL), L’Hospitalet de Llobregat, Barcelona, Spain; Consortium for Biomedical Research in Epidemiology and Public Health (CIBERESP), Madrid, Spain; Department of Clinical Sciences, Faculty of Medicine, University of Barcelona, Barcelona, Spain; University Hospitals Bristol and Weston NHS Foundation Trust, National Institute for Health Research Bristol; Biomedical Research Centre, University of Bristol, Bristol, UK

**Author notes:** **Corresponding author:** James Yarmolinsky, PhD, MRC Integrative Epidemiology Unit, Population Health Sciences, University of Bristol, Bristol, UK.

## Abstract

**Background:** Preclinical and human genetics studies suggest that impaired glucose-dependent insulinotropic polypeptide receptor (GIPR) signalling worsens glycaemic control. The relationship between GIPR signalling and risk of cancers influenced by impaired glucose homeostasis is unclear.

**Methods:** We examined the association of a missense variant in *GIPR*, rs1800437 (E354Q), shown to impair long-term GIPR signalling and lower circulating GIP concentrations, with risk of 6 cancers influenced by impaired glucose homeostasis. Summary genetic association data on breast, colorectal, endometrial, lung, pancreatic, and renal cancer risk were obtained from GWAS consortia (235,698 cases, 333,932 controls). Replication analyses were performed in the FinnGen consortium (8,401 breast cancer cases, 99,321 controls). Colocalisation was performed to examine robustness of findings to genetic confounding. Finally, we examined the association of E354Q with potential downstream molecular mediators of GIPR signalling to identify possible mechanisms underpinning an effect on cancer risk.

**Results:** Each copy of E354Q was associated with a higher risk of overall breast cancer (OR:1.05,95%CI:1.03-1.06,*P*=4.23×10^−9^) and luminal A-like breast cancer (OR:1.05,95%CI:1.03-1.06,*P*=6.02×10^−7^). In replication analysis, E354Q was likewise associated with breast cancer risk (OR:1.06,95%CI:1.02-1.08,*P*=1.09×10^−3^) and in colocalisation there was a >99.9% posterior probability of circulating GIP concentrations sharing a causal variant with overall and luminal A-like breast cancer in *GIPR*. E354Q was not associated with risk of the 5 other cancers examined. In mechanistic analysis, this variant was associated with higher postprandial glucose concentrations but diminished insulin secretion and lower testosterone concentrations.

**Conclusion:** Our comprehensive drug target-Mendelian randomization analysis across 6 cancers suggests an adverse effect of the *GIPR* E354Q variant on overall and luminal A-like breast cancer risk. These findings provide genetic evidence to support further evaluation of the therapeutic potential of GIPR signalling in breast cancer prevention.

---

Preclinical and epidemiological studies suggest an important role of dysregulated metabolism in cancer development, in particular carcinogenic effects of sustained elevated insulin levels^1,2^. Hyperinsulinaemia has consistently been associated with risk of several cancers in both observational and genetic epidemiological studies^3–9^. *In vitro* studies have demonstrated that insulin signalling is mitogenic on cancer cells and can induce cell migration, providing possible mechanisms for carcinogenesis^10^. Enhanced understanding of molecular mechanisms regulating insulin signalling could inform the development of potential therapeutic strategies for cancer prevention.

Glucose-dependent insulinotropic peptide (GIP) is one of two incretin hormones, along with glucagon-like peptide-1 (GLP1), that are produced in response to nutrient consumption, maintaining glucose homeostasis by increasing insulin and lowering glucagon secretion^11^. In a phase 3 clinical trial, tirzepatide, a dual GIPR/GLP1R agonist, was shown to confer superior HbA_1c_ control as compared to GLP1R agonism alone and has recently been approved by the U.S. Food and Drug Administration (FDA) for type 2 diabetes treatment^12,13^. By potentiating postprandial insulin secretion and increasing blood insulin levels, there is some concern that pharmacological agonism of the GIPR signalling pathway could increase risk of hyperinsulinemia-driven cancers^14^. However, the few epidemiological studies that have examined the relationship between circulating GIP concentrations and cancer risk have generated conflicting results^15–17^.

Naturally occurring variation in genes encoding drug targets can be leveraged to predict the effect of pharmacological perturbation of these targets on disease risk (“drug-target Mendelian randomization”)^18^. Since germline genetic variants are randomly assorted at meiosis and fixed at conception, such studies should be less prone to confounding than conventional observational studies and cannot be influenced by reverse causation^19,20^. In addition, drug-target Mendelian randomization permits the effect of the long-term perturbation of drug targets on cancer risk to be examined. This is advantageous when evaluating cancer outcomes given long induction periods for cancer development and the number of emerging drugs that do not have long-term efficacy data^20,21^.

Here, we used a missense variant in *GIPR*, previously shown to result in impaired long-term GIPR signalling and decreased fasting and 2-hour GIP concentrations, to predict the potential effect of such impaired GIPR signalling on risk of 6 cancers influenced by hyperinsulinemia (overall and histotype-specific breast, colorectal, endometrial, lung, pancreatic, and renal cancers)^22,23^. We tested findings for replication in the Finngen Consortium and employed colocalisation to evaluate their robustness to violations of Mendelian randomization assumptions. Finally, we used this variant to examine potential downstream mediators of GIPR signalling (i.e. various measures of childhood and adult adiposity, fasting and postprandial glucose and insulin, endogenous sex hormones, and lipids), to identify possible mechanisms underpinning an effect of impaired GIPR signalling on cancer risk.

## Methods

### Study population

Summary genetic association data on overall and histological subtype-specific cancer susceptibility were obtained from genome-wide association study (GWAS) meta-analyses of 6 adult cancers in up to 235,698 cases and 333,932 of European ancestry. Cancer sites were selected based on previous genetic epidemiological evidence linking fasting insulin to cancer susceptibility and included the following anatomical sites: breast (133,384 cases, 113,789 controls), colorectum (58,221 cases, 67,694 controls), endometrium (12,906 cases, 108,979 controls), lung (11,348 cases, 15,861 controls), kidney (10,784 cases, 20,406 controls), and pancreas (9,055 cases, 7,203 controls)^3–5,7–9,24–29^. Further information on numbers of cases and controls across histological subtype-stratified analyses is presented in **Table 2**.

For replication analyses, summary genetic association data were obtained on 8,401 breast cancer cases and 99,321 controls of European ancestry from the Finngen consortium^23^. We also performed exploratory analyses examining the association of impaired GIPR signalling with breast cancer risk in *BRCA1/2* mutation carriers, by obtaining GWAS summary data on 19,036 *BRCA1* carriers, 12,412 *BRCA2* carriers and 100,594 controls of European ancestry from the Consortium of Investigations of Modifiers of BRCA1/2 (CIMBA)^31,32^.

For analyses investigating the effect of impaired GIPR signalling on putative mediators of the GIPR-breast cancer relationship, we obtained summary genetic association data from previous GWAS of child and adult BMI or self-reported comparative body size, type 2 diabetes, 3 endogenous sex hormones, 4 glycaemic traits measured in the non-postprandial state, 10 glycaemic traits measured following an oral glucose tolerance test, 2 lipid traits, and insulin-like growth factor 1^33–43^. These traits were selected based on previous observational and genetic epidemiological evidence supporting their potential role in breast cancer risk^38,44–48^. All 14 glycaemic traits were measured in non-diabetic individuals. Additional information on the specific traits included, their measurement, along with participant characteristics and covariates included in adjustment strategies across each GWAS are presented in **Supplementary Table 1**. Further information on imputation, statistical analyses and quality control measures for these studies can be found in the original publications.

### Instrument construction

We used a missense variant in *GIPR*, rs1800437 (E354Q, C allele), to proxy impaired GIPR signalling. This variant has been implicated in increased GIP residence time at GIPR, increased internalisation and signalling, and thus desensitisation and impairment of the signalling pathway long-term^23^. This variant was also associated (*P*<5.0×10^−8^) with lower fasting and 2-hour GIP concentrations in a GWAS meta-analysis of 7,828 individuals of European ancestry across the Malmö Diet and Cancer (MDC) and Prevalence, Prediction and Prevention of diabetes (PPP)-Botnia studies. Participants in both studies were not taking anti-diabetic medications^22^.

To generate genetic instruments to proxy potential mediators of the GIPR signalling-cancer relationship, genome-wide significant (*P*<5.0×10^−8^) and independent (r^2^<0.001) SNPs were selected using the 1000 Genomes Phase 3 European reference panel^49^.

### Statistical analysis

Analyses of the effect of traits influenced by E354Q on cancer risk (i.e. putative mediators of the effect of E354Q on cancer risk) were performed using inverse-variance weighted (IVW) random-effects models^50^.

Mendelian randomization (MR) analysis assumes that a genetic instrument (i) is associated with a modifiable exposure or drug target (“relevance”), (ii) does not share a common cause with an outcome (“exchangeability”), and (iii) has no direct effect on the outcome (“exclusion restriction”) ^51,52^. Under the assumption of monotonicity (i.e. the direction of effect of the instrument on the exposure is consistent across all individuals), MR can provide valid point estimates for those participants whose exposure is influenced by the instrument (i.e. a local average treatment effect^53^).

We assessed the “relevance” assumption by generating estimates of the proportion of variance in each trait explained by the instrument (r^2^) and F-statistics. An F-statistic >10 is conventionally used to indicate that instruments are unlikely to suffer from weak instrument bias^54^.

Colocalisation was performed as a sensitivity analysis for primary analyses where there was nominal evidence of an association (*P*<0.05), to assess whether two traits examined shared a causal variant at a genetic locus (e.g. as opposed to both traits having distinct causal variants that are in linkage disequilibrium)^55^. Colocalisation analyses were performed using the coloc R package by generating ±250kb windows from the sentinel SNP used to proxy the instrument^55^. We used H4>50.0% as evidence to support colocalisation of traits.

When testing the effect of putative GIPR signalling-cancer mediators on cancer risk, we evaluated the “exclusion restriction” assumption through performing various sensitivity analyses, including MR-Egger, weighted median, and weighted mode estimation^51–53^. We also performed iterative “leave-one-out” analysis to examine the robustness of findings to individual influential SNPs in IVW models.

To account for multiple testing across E354Q-cancer analyses, a Bonferroni correction was used to establish a *P*-value threshold of <0.0029 (false positive rate=0.05/17 statistical tests, representing 17 cancer endpoints), which we used as a heuristic to define “strong evidence,” with findings between *P*≥0.0029 and *P*<0.05 defined as “weak evidence.”

## Results

Characteristics of genetic variants used to proxy all traits are presented in **Supplementary Table 2**. F-statistics for genetic instruments for these traits ranged from 57.7 to 30,028.7, suggesting that our analyses were unlikely to suffer from weak instrument bias (**Table 1**).

**Table 1.** Instrument strength estimates across all traits examined.

| Trait (units) | N of SNPs | R<sup>2</sup> | F-stats |
| --- | --- | --- | --- |
| Bioavailable testosterone (ln-transformed, nmol/L) | 178 | 0.054 | 10,760.2 |
| Total testosterone (inverse normal rank transformed, nmol/L) | 256 | 0.074 | 18,454.2 |
| Type 2 diabetes (BMI adj.) | 54 | 0.017 | 5,241.8 |
| 2 hour glucose (mmol⋅L<sup>-1</sup>) | 12 | 0.0028 | 790.3 |
| HbA<sub>1c</sub> (%) | 64 | 0.026 | 7,552.5 |
| BMI (sex-combined) (SD) | 419 | 0.061 | 30,028.7 |
| BMI (female) (SD) | 36 | 0.014 | 2,463.7 |
| Comparative body size at age 10 (SD) | 209 | 0.035 | 16,720.0 |

R<sup>2</sup> is an estimate of the proportion of variance in each trait explained by the instrument. An F-statistic >10 is conventionally used to indicate that instruments are unlikely to suffer from weak instrument bias<sup>54</sup>. In analyses of the effect of E354Q on breast cancer risk scaled to the effect of this variant on GIP concentrations, r<sup>2</sup> and F-statistics for fasting and 2-hour GIP concentrations were: 0.0073 and 57.7, 0.0085 and 64.0, respectively. Summary genetic association data on fasting and 2-hour GIP concentrations from Almgren et al were obtained from the MDC subcohort because of denser variant coverage as compared to the PPP-Botnia study. HbA<sub>1c</sub> = glycated haemoglobin, BMI = body mass index (adult), comparative body size at age 10 = recall of an individual's body size at age 10 as compared to average.

**Table 2.** Association between E354Q and overall and histotype-specific breast, endometrial, colorectal, lung, renal, and pancreatic cancer risk.

| Outcome | No. of cases | No. of controls | OR (95% CI) | P value |
| --- | --- | --- | --- | --- |
| Breast cancer | 133,384 | 113,789 | 1.05 (1.03, 1.06) | $6.26 \times 10^{-9}$ |
| Luminal A-like | N/A* | N/A* | 1.05 (1.03, 1.06) | $6.02 \times 10^{-7}$ |
| Luminal B-like | N/A* | N/A* | 1.04 (0.99, 1.09) | 0.09 |
| Luminal B-HER2-Negative-Like | N/A* | N/A* | 1.06 (1.02, 1.08) | $1.82 \times 10^{-3}$ |
| HER2-enriched-like | N/A* | N/A* | 1.05 (0.99, 1.11) | 0.11 |
| Triple-negative | N/A* | N/A* | 1.01 (0.97, 1.08) | 0.29 |
| Colorectal cancer | 58,221 | 67,694 | 1.01 (0.98, 1.06) | 0.63 |
| Colon cancer | 32,002 | 64,159 | 0.93 (0.90, 1.07) | 0.81 |
| Rectal cancer | 16,212 | 64,159 | 1.01 (0.98, 1.08) | 0.59 |
| Endometrial cancer | 12,906 | 108,979 | 0.97 (0.93, 1.08) | 0.09 |
| Endometrioid Endometrial cancer | 8,758 | 108,979 | 0.98 (0.94, 1.09) | 0.35 |
| Non-Endometrioid Endometrial cancer | 1,230 | 108,979 | 0.96 (0.86, 1.16) | 0.46 |
| Lung cancer | 11,348 | 15,861 | 1.01 (0.97, 1.09) | 0.61 |
| Lung adenocarcinoma | 3,442 | 14,894 | 1.05 (0.98, 1.11) | 0.17 |
| Squamous cell carcinoma | 3,275 | 15,038 | 0.98 (0.92, 1.11) | 0.57 |
| Renal cancer | 10,784 | 20,406 | 0.95 (0.91, 0.99) | 0.01 |
| Pancreatic cancer | 9,055 | 7,203 | 0.97 (0.92, 1.10) | 0.28 |

OR represents the exponential increase in odds per copy of E354Q (rs1800437, C allele). \*Effective sample size not available.

### Association of E354Q with cancer risk

Each copy of E354Q was strongly associated with a higher risk of breast cancer (OR:1.05, 95%CI:1.03-1.06, *P*=6.26×10^−9^)(**Table 2**). In histological subtype-stratified analyses, E354Q was also strongly associated with higher risk of luminal A-like (OR:1.05, 95%CI:1.03-1.06, *P*=6.02×10^−7^) and luminal B HER2 negative-like breast cancer (OR:1.06, 95%CI:1.02-1.09, *P*=1.82×10^−3^)(**Table 2**). When scaled to a 1 unit lowering of ln-fasting GIP concentrations mediated by this variant this represents ORs (95%CIs) of 1.82 (1.47-2.17), 1.92 (1.49-2.50), and 2.17 (1.33-3.57) for overall, luminal A-like, and luminal B HER2 negative-like breast cancer, respectively. Colocalisation analysis suggested that fasting and 2-hour GIP concentrations had a >99.9% posterior probability of sharing a causal variant with both overall and luminal A-like breast cancer risk within the *GIPR* locus and a >51.8% probability of sharing a causal variant with luminal B HER2 negative-like breast cancer (**Table 3)**.

**Table 3.** Colocalisation analysis results for fasting and 2-hour GIP concentrations and cancer risk in the *GIPR* locus.

| Exposure | Outcome | H<sub>0</sub> | H<sub>1</sub> | H<sub>2</sub> | H<sub>3</sub> | H<sub>4</sub> |
| --- | --- | --- | --- | --- | --- | --- |
| Fasting GIP | Overall breast cancer | $1.84 \times 10^{-6}$ | $1.19 \times 10^{-4}$ | $3.21 \times 10^{-4}$ | $1.07 \times 10^{-3}$ | $9.99 \times 10^{-1}$ |
| Fasting GIP | Luminal A | $1.36 \times 10^{-4}$ | $8.76 \times 10^{-4}$ | $3.24 \times 10^{-4}$ | $1.09 \times 10^{-3}$ | $9.98 \times 10^{-1}$ |
| Fasting GIP | Luminal B HER2 Negative | $6.43 \times 10^{-2}$ | $4.15 \times 10^{-1}$ | $4.10 \times 10^{-4}$ | $2.13 \times 10^{-3}$ | $5.19 \times 10^{-1}$ |
| Fasting GIP | Renal cancer | $1.09 \times 10^{-1}$ | $7.03 \times 10^{-1}$ | $1.02 \times 10^{-3}$ | $6.36 \times 10^{-3}$ | $1.80 \times 10^{-1}$ |
| 2-hour GIP | Overall breast cancer | $9.14 \times 10^{-7}$ | $1.25 \times 10^{-5}$ | $1.59 \times 10^{-4}$ | $1.18 \times 10^{-3}$ | $9.99 \times 10^{-1}$ |
| 2-hour GIP | Luminal A | $6.68 \times 10^{-5}$ | $9.14 \times 10^{-4}$ | $1.59 \times 10^{-4}$ | $1.18 \times 10^{-3}$ | $9.98 \times 10^{-1}$ |
| 2-hour GIP | Luminal B HER2 Negative | $3.17 \times 10^{-2}$ | $4.61 \times 10^{-1}$ | $9.68 \times 10^{-14}$ | $2.23 \times 10^{-3}$ | $5.32 \times 10^{-1}$ |
| 2-hour GIP | Renal cancer | $5.33 \times 10^{-2}$ | $7.29 \times 10^{-1}$ | $4.95 \times 10^{-4}$ | $6.56 \times 10^{-3}$ | $2.11 \times 10^{-1}$ |

H<sub>0</sub>-H<sub>4</sub>: posterior probabilities of the associations between the 2 traits examined, evaluating 5 different configurations:
- H<sub>0</sub>: Neither trait has an association in the region - H<sub>1</sub>: The first trait has an association in the region but the second does not - H<sub>2</sub>: The second trait has an association in the region but the first does not - H<sub>3</sub>: Both traits have an association in the region but these SNPs are independent - H<sub>4</sub>: Both traits have an association in the region and share a SNP

In analyses across five other cancer sites, there was weak evidence for an association of E354Q with lower risk of renal cancer (OR:0.95, 95%CI:0.91-0.99, *P*=0.01), but little evidence of association of this variant with risk of 5 other cancers examined (**Tables 2,3**). In colocalisation analysis there was little evidence to support one or more shared causal variants for fasting or 2-hour GIP concentrations and renal cancer risk in *GIPR* (H4<21.2%; **Table 3**).

### Replication analyses in Finngen and exploratory analyses in *BRCA1/2* mutation carriers

Findings for an association of E354Q with breast cancer risk were replicated in an independent sample of 8,401 cases and 99,321 controls in the FinnGen consortium (OR:1.06, 95%CI:1.02-1.08, *P*=1.09×10^−3^). In exploratory analyses in 31,448 cases and 100,594 controls with *BRCA1* or *BRCA2* mutations, there was little evidence of association of E354Q with breast cancer risk (*BRCA1*:OR 1.00, 95%CI:0.96-1.09, *P*=0.98; *BRCA2*:OR:1.04, 95%CI:0.98-1.10, *P*=0.16).

### Type 2 diabetes, body mass index, glycaemic traits, lipids and sex hormones as potential mediators of an association of E354Q with breast cancer risk

In combined Mendelian randomization and colocalisation analyses, we found consistent evidence to implicate E354Q in higher risk of type 2 diabetes (BMI adj.)(OR:1.06, 95%CI:1.04-1.06, *P*=6.80×10^−12^; H_4_≥90.0%) and lower BMI (-0.034SD change, 95%CI:-0.039,-0.029, *P*=7.08×10^−42^, H_4_ =99.9%). The association of E354Q with BMI was consistent in sensitivity analyses using female-specific BMI association estimates (-0.032SD change, 95%CI:-0.042,-0.022, *P*=5.79×10^−42^, H_4_ =99.8%) (**Tables 4, 6**). We also found consistent evidence to implicate E354Q in lower comparative body size aged 10 (-0.012SD change, 95%CI:-0.015,-0.0083, *P*=3.10×10^11^, H_4_ =99.9%), although there was no evidence for an association with measured BMI in children aged 2-10 (0.0014SD change, 95% CI:-0.018,0.021, *P*=0.89) (**Tables 4, 6**).

**Table 4.**
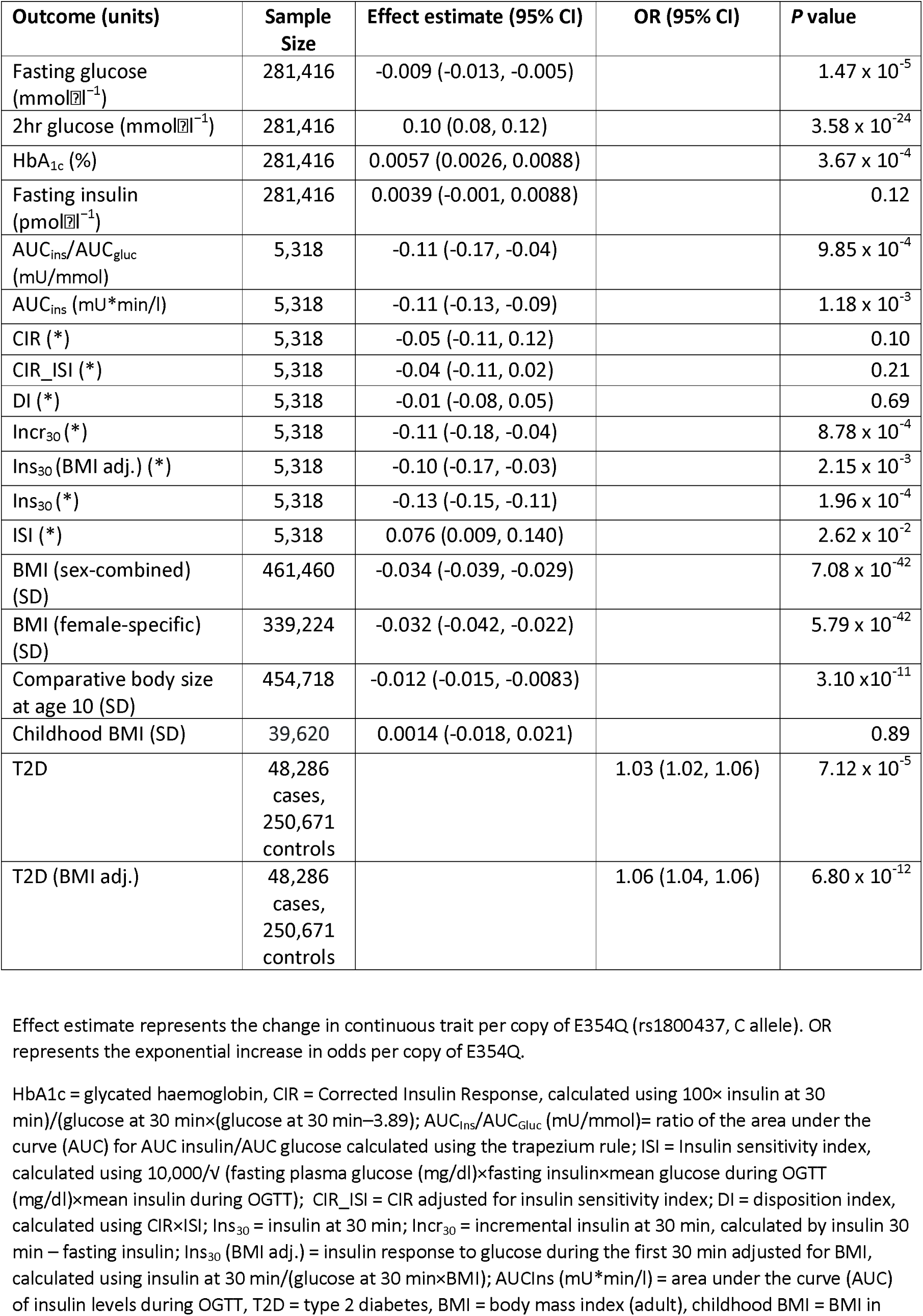
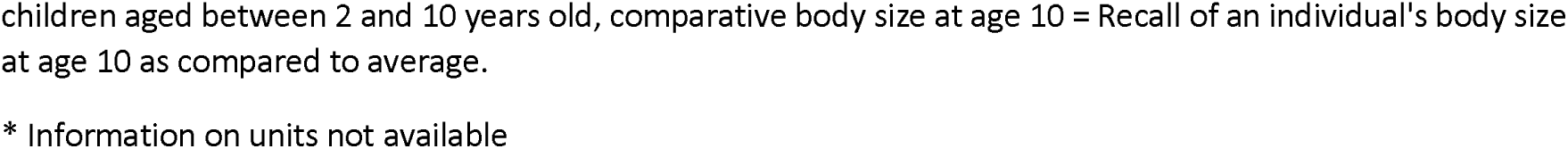
Association between E354Q and glycaemic traits, adiposity measures, and type 2 diabetes (adjusted and unadjusted for BMI).

| Outcome (units) | Sample Size | Effect estimate (95% CI) | OR (95% CI) | P value |
| --- | --- | --- | --- | --- |
| Fasting glucose (mmol l<sup>-1</sup>) | 281,416 | -0.009 (-0.013, -0.005) |  | 1.47 x 10<sup>-5</sup> |
| 2hr glucose (mmol l<sup>-1</sup>) | 281,416 | 0.10 (0.08, 0.12) |  | 3.58 x 10<sup>-24</sup> |
| HbA<sub>1c</sub> (%) | 281,416 | 0.0057 (0.0026, 0.0088) |  | 3.67 x 10<sup>-4</sup> |
| Fasting insulin (pmol l<sup>-1</sup>) | 281,416 | 0.0039 (-0.001, 0.0088) |  | 0.12 |
| AUC<sub>ins</sub>/AUC<sub>gluc</sub> (mU/mmol) | 5,318 | -0.11 (-0.17, -0.04) |  | 9.85 x 10<sup>-4</sup> |
| AUC<sub>ins</sub> (mU*min/l) | 5,318 | -0.11 (-0.13, -0.09) |  | 1.18 x 10<sup>-3</sup> |
| CIR (*) | 5,318 | -0.05 (-0.11, 0.12) |  | 0.10 |
| CIR_ISI (*) | 5,318 | -0.04 (-0.11, 0.02) |  | 0.21 |
| DI (*) | 5,318 | -0.01 (-0.08, 0.05) |  | 0.69 |
| Incr<sub>30</sub> (*) | 5,318 | -0.11 (-0.18, -0.04) |  | 8.78 x 10<sup>-4</sup> |
| Ins<sub>30</sub> (BMI adj.) (*) | 5,318 | -0.10 (-0.17, -0.03) |  | 2.15 x 10<sup>-3</sup> |
| Ins<sub>30</sub> (*) | 5,318 | -0.13 (-0.15, -0.11) |  | 1.96 x 10<sup>-4</sup> |
| ISI (*) | 5,318 | 0.076 (0.009, 0.140) |  | 2.62 x 10<sup>-2</sup> |
| BMI (sex-combined) (SD) | 461,460 | -0.034 (-0.039, -0.029) |  | 7.08 x 10<sup>-42</sup> |
| BMI (female-specific) (SD) | 339,224 | -0.032 (-0.042, -0.022) |  | 5.79 x 10<sup>-42</sup> |
| Comparative body size at age 10 (SD) | 454,718 | -0.012 (-0.015, -0.0083) |  | 3.10 x 10<sup>-11</sup> |
| Childhood BMI (SD) | 39,620 | 0.0014 (-0.018, 0.021) |  | 0.89 |
| T2D | 48,286 cases, 250,671 controls |  | 1.03 (1.02, 1.06) | 7.12 x 10<sup>-5</sup> |
| T2D (BMI adj.) | 48,286 cases, 250,671 controls |  | 1.06 (1.04, 1.06) | 6.80 x 10<sup>-12</sup> |

Effect estimate represents the change in continuous trait per copy of E354Q (rs1800437, C allele). OR represents the exponential increase in odds per copy of E354Q.
HbA<sub>1c</sub> = glycated haemoglobin, CIR = Corrected Insulin Response, calculated using 100× insulin at 30 min)/(glucose at 30 min×(glucose at 30 min–3.89); AUC<sub>ins</sub>/AUC<sub>gluc</sub> (mU/mmol)= ratio of the area under the curve (AUC) for AUC insulin/AUC glucose calculated using the trapezium rule; ISI = Insulin sensitivity index, calculated using 10,000/V (fasting plasma glucose (mg/dl)×fasting insulin×mean glucose during OGTT (mg/dl)×mean insulin during OGTT); CIR\_ISI = CIR adjusted for insulin sensitivity index; DI = disposition index, calculated using CIR×ISI; Ins<sub>30</sub> = insulin at 30 min; Incr<sub>30</sub> = incremental insulin at 30 min, calculated by insulin 30 min – fasting insulin; Ins<sub>30</sub> (BMI adj.) = insulin response to glucose during the first 30 min adjusted for BMI, calculated using insulin at 30 min/(glucose at 30 min×BMI); AUC<sub>ins</sub> (mU\*min/l) = area under the curve (AUC) of insulin levels during OGTT, T2D = type 2 diabetes, BMI = body mass index (adult), childhood BMI = BMI in
children aged between 2 and 10 years old, comparative body size at age 10 = Recall of an individual's body size at age 10 as compared to average.
\* Information on units not available

**Table 5.**
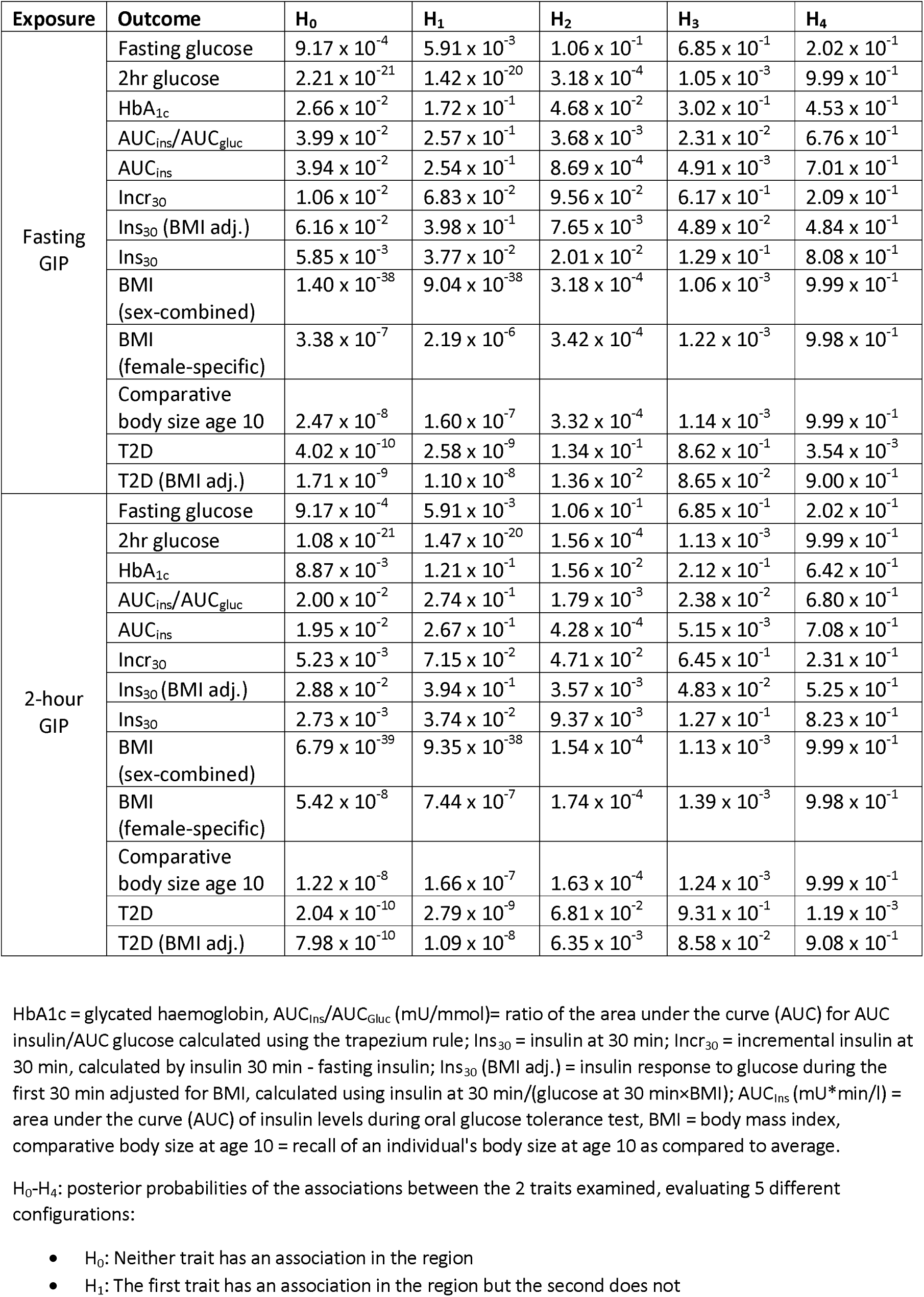
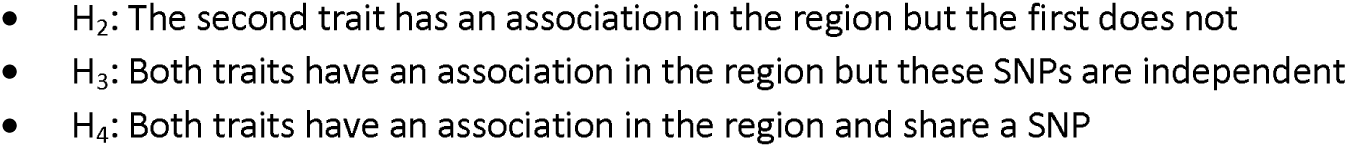
Colocalisation analysis results for fasting and 2-hour GIP concentrations, BMI (sex-combined and female-specific) and type 2 diabetes and glycaemic traits in the *GIPR* locus.

| Exposure | Outcome | H<sub>0</sub> | H<sub>1</sub> | H<sub>2</sub> | H<sub>3</sub> | H<sub>4</sub> |
| --- | --- | --- | --- | --- | --- | --- |
| Fasting GIP | Fasting glucose | $9.17 \times 10^{-4}$ | $5.91 \times 10^{-3}$ | $1.06 \times 10^{-1}$ | $6.85 \times 10^{-1}$ | $2.02 \times 10^{-1}$ |
| | 2hr glucose | $2.21 \times 10^{-21}$ | $1.42 \times 10^{-20}$ | $3.18 \times 10^{-4}$ | $1.05 \times 10^{-3}$ | $9.99 \times 10^{-1}$ |
| | HbA<sub>1c</sub> | $2.66 \times 10^{-2}$ | $1.72 \times 10^{-1}$ | $4.68 \times 10^{-2}$ | $3.02 \times 10^{-1}$ | $4.53 \times 10^{-1}$ |
| | AUC<sub>ins</sub>/AUC<sub>gluc</sub> | $3.99 \times 10^{-2}$ | $2.57 \times 10^{-1}$ | $3.68 \times 10^{-3}$ | $2.31 \times 10^{-2}$ | $6.76 \times 10^{-1}$ |
| | AUC<sub>ins</sub> | $3.94 \times 10^{-2}$ | $2.54 \times 10^{-1}$ | $8.69 \times 10^{-4}$ | $4.91 \times 10^{-3}$ | $7.01 \times 10^{-1}$ |
| | Incr<sub>30</sub> | $1.06 \times 10^{-2}$ | $6.83 \times 10^{-2}$ | $9.56 \times 10^{-2}$ | $6.17 \times 10^{-1}$ | $2.09 \times 10^{-1}$ |
| | Ins<sub>30</sub> (BMI adj.) | $6.16 \times 10^{-2}$ | $3.98 \times 10^{-1}$ | $7.65 \times 10^{-3}$ | $4.89 \times 10^{-2}$ | $4.84 \times 10^{-1}$ |
| | Ins<sub>30</sub> | $5.85 \times 10^{-3}$ | $3.77 \times 10^{-2}$ | $2.01 \times 10^{-2}$ | $1.29 \times 10^{-1}$ | $8.08 \times 10^{-1}$ |
| | BMI (sex-combined) | $1.40 \times 10^{-38}$ | $9.04 \times 10^{-38}$ | $3.18 \times 10^{-4}$ | $1.06 \times 10^{-3}$ | $9.99 \times 10^{-1}$ |
| | BMI (female-specific) | $3.38 \times 10^{-7}$ | $2.19 \times 10^{-6}$ | $3.42 \times 10^{-4}$ | $1.22 \times 10^{-3}$ | $9.98 \times 10^{-1}$ |
| | Comparative body size age 10 | $2.47 \times 10^{-8}$ | $1.60 \times 10^{-7}$ | $3.32 \times 10^{-4}$ | $1.14 \times 10^{-3}$ | $9.99 \times 10^{-1}$ |
| | T2D | $4.02 \times 10^{-10}$ | $2.58 \times 10^{-9}$ | $1.34 \times 10^{-1}$ | $8.62 \times 10^{-1}$ | $3.54 \times 10^{-3}$ |
| | T2D (BMI adj.) | $1.71 \times 10^{-9}$ | $1.10 \times 10^{-8}$ | $1.36 \times 10^{-2}$ | $8.65 \times 10^{-2}$ | $9.00 \times 10^{-1}$ |
| 2-hour GIP | Fasting glucose | $9.17 \times 10^{-4}$ | $5.91 \times 10^{-3}$ | $1.06 \times 10^{-1}$ | $6.85 \times 10^{-1}$ | $2.02 \times 10^{-1}$ |
| | 2hr glucose | $1.08 \times 10^{-21}$ | $1.47 \times 10^{-20}$ | $1.56 \times 10^{-4}$ | $1.13 \times 10^{-3}$ | $9.99 \times 10^{-1}$ |
| | HbA<sub>1c</sub> | $8.87 \times 10^{-3}$ | $1.21 \times 10^{-1}$ | $1.56 \times 10^{-2}$ | $2.12 \times 10^{-1}$ | $6.42 \times 10^{-1}$ |
| | AUC<sub>ins</sub>/AUC<sub>gluc</sub> | $2.00 \times 10^{-2}$ | $2.74 \times 10^{-1}$ | $1.79 \times 10^{-3}$ | $2.38 \times 10^{-2}$ | $6.80 \times 10^{-1}$ |
| | AUC<sub>ins</sub> | $1.95 \times 10^{-2}$ | $2.67 \times 10^{-1}$ | $4.28 \times 10^{-4}$ | $5.15 \times 10^{-3}$ | $7.08 \times 10^{-1}$ |
| | Incr<sub>30</sub> | $5.23 \times 10^{-3}$ | $7.15 \times 10^{-2}$ | $4.71 \times 10^{-2}$ | $6.45 \times 10^{-1}$ | $2.31 \times 10^{-1}$ |
| | Ins<sub>30</sub> (BMI adj.) | $2.88 \times 10^{-2}$ | $3.94 \times 10^{-1}$ | $3.57 \times 10^{-3}$ | $4.83 \times 10^{-2}$ | $5.25 \times 10^{-1}$ |
| | Ins<sub>30</sub> | $2.73 \times 10^{-3}$ | $3.74 \times 10^{-2}$ | $9.37 \times 10^{-3}$ | $1.27 \times 10^{-1}$ | $8.23 \times 10^{-1}$ |
| | BMI (sex-combined) | $6.79 \times 10^{-39}$ | $9.35 \times 10^{-38}$ | $1.54 \times 10^{-4}$ | $1.13 \times 10^{-3}$ | $9.99 \times 10^{-1}$ |
| | BMI (female-specific) | $5.42 \times 10^{-8}$ | $7.44 \times 10^{-7}$ | $1.74 \times 10^{-4}$ | $1.39 \times 10^{-3}$ | $9.98 \times 10^{-1}$ |
| | Comparative body size age 10 | $1.22 \times 10^{-8}$ | $1.66 \times 10^{-7}$ | $1.63 \times 10^{-4}$ | $1.24 \times 10^{-3}$ | $9.99 \times 10^{-1}$ |
| | T2D | $2.04 \times 10^{-10}$ | $2.79 \times 10^{-9}$ | $6.81 \times 10^{-2}$ | $9.31 \times 10^{-1}$ | $1.19 \times 10^{-3}$ |
| | T2D (BMI adj.) | $7.98 \times 10^{-10}$ | $1.09 \times 10^{-8}$ | $6.35 \times 10^{-3}$ | $8.58 \times 10^{-2}$ | $9.08 \times 10^{-1}$ |

HbA<sub>1c</sub> = glycated haemoglobin, AUC<sub>ins</sub>/AUC<sub>gluc</sub> (mU/mmol) = ratio of the area under the curve (AUC) for AUC insulin/AUC glucose calculated using the trapezium rule; Ins<sub>30</sub> = insulin at 30 min; Incr<sub>30</sub> = incremental insulin at 30 min, calculated by insulin 30 min - fasting insulin; Ins<sub>30</sub> (BMI adj.) = insulin response to glucose during the first 30 min adjusted for BMI, calculated using insulin at 30 min/(glucose at 30 min×BMI); AUC<sub>ins</sub> (mU\*min/l) = area under the curve (AUC) of insulin levels during oral glucose tolerance test, BMI = body mass index, comparative body size at age 10 = recall of an individual's body size at age 10 as compared to average.
H<sub>0</sub>-H<sub>4</sub>: posterior probabilities of the associations between the 2 traits examined, evaluating 5 different configurations:
- H<sub>0</sub>: Neither trait has an association in the region - H<sub>1</sub>: The first trait has an association in the region but the second does not
- $H_2$ : The second trait has an association in the region but the first does not - $H_3$ : Both traits have an association in the region but these SNPs are independent - $H_4$ : Both traits have an association in the region and share a SNP

**Table 6:** Association between E354Q and sex hormone measures, lipid measures, HOMA-IR, and IGF-1.

| Outcome (units) | Sample size | Effect estimate (95% CI) | P value | Data Source |
| --- | --- | --- | --- | --- |
| SHBG (BMI adj.) (ln-transformed, nmol/L) | 188,908 | -0.0037 (-0.0068, -0.00067) | 0.0037 | <sup>38</sup> |
| SHBG (inverse normal rank transformed, nmol/L) | 189,473 | 0.0040 (0.00049, 0.00075) | 0.098 | <sup>38</sup> |
| Bioavailable testosterone (ln-transformed, nmol/L) | 188,507 | -0.019 (-0.025, -0.012) | $5.20 \times 10^{-9}$ | <sup>38</sup> |
| Total testosterone (inverse normal rank transformed, nmol/L) | 230,454 | -0.022 (-0.029, -0.015) | $5.00 \times 10^{-10}$ | <sup>38</sup> |
| HDL-c (mg/dL) | 1,320,000 | -0.013 (-0.016, -0.0095) | $4.59 \times 10^{-4}$ | <sup>43</sup> |
| LDL-c (mg/dL) | 1,320,000 | -0.020 (-0.023, -0.016) | $2.83 \times 10^{-30}$ | <sup>43</sup> |
| HOMA-IR | 37,037 | -0.0057 (-0.016, 0.0051) | 0.30 | <sup>41</sup> |
| IGF-1 (nmol/L) | 342,439 | 0.0088 (0.0030, 0.015) | $2.75 \times 10^{-3}$ | <sup>42</sup> |

BMI = body mass index, SHBG = sex hormone-binding globulin, HDL = high density lipoprotein, LDL = low density lipoprotein, HOMA-IR = Homeostatic measure of insulin resistance, IGF-1 = Insulin-like growth factor 1.

Each copy of E354Q was also associated with higher 2-hour glucose concentrations (0.10, 95%CI:0.08-0.12, *P*=3.58×10^−24^, H_4_ =99.9%) and lower levels of 3 measures of insulin secretion: AUC _ins_ (-0.11, 95%CI:-0.13,-0.09, *P*=1.18×10^−3^, H_4_ ≥70.1%), AUC _ins_ /AUC_gluc_ (-0.11, 95%CI:-0.17,-0.04, *P*=9.85×10^−4^, H_4_ ≥67.6%), and Ins_30_ (-0.13, 95%CI:-0.15,-0.11, *P*=1.96×10^−4^, H_4_ ≥80.8%)**(Tables 4,5)**. Evidence of an association of E354Q with HbA_1c_ (0.0057% change, 95%CI:0.0026,0.0088, *P*=3.67×10^−4^) and Ins_30_ (BMI adj.)(-0.10, 95%CI:-0.17,-0.03, *P*=2.15×10^−3^) was also supported in colocalisation analysis for 2-hour, but not fasting, GIP concentrations (H_4_= 64.2% and 52.4% for HbA_1c_ and Ins30 (BMI adj.), respectively)(**Tables 4,5**).

When examining hormone and lipid traits, there was also consistent MR and colocalisation evidence to implicate E354Q in lower total (-0.022, 95%CI:-0.029,-0.015, *P*=5.00×10^−10^, H_4_ =99.8%) and bioavailable testosterone concentrations (-0.019, 95%CI:-0.025,-0.012, *P*=5.20×10^−9^, H_4_ ≥99.5%). Full MR and colocalisation estimates across all potential mediators examined are presented in **Tables 4-7**. Findings from iterative leave-one-out analysis are presented in **Supplementary Table 3**.

**Table 7.** Colocalisation analysis results for fasting and 2-hour GIP concentrations and sex hormone measures, lipid measures and IGF-1.

| Exposure | Outcome | H<sub>0</sub> | H<sub>1</sub> | H<sub>2</sub> | H<sub>3</sub> | H<sub>4</sub> |
| --- | --- | --- | --- | --- | --- | --- |
| Fasting GIP | SHBG (BMI adj.) | $8.75 \times 10^{-41}$ | $5.65 \times 10^{-40}$ | $1.32 \times 10^{-1}$ | $8.50 \times 10^{-1}$ | $1.80 \times 10^{-2}$ |
| | Bioavailable testosterone | $2.53 \times 10^{-6}$ | $1.63 \times 10^{-5}$ | $8.05 \times 10^{-4}$ | $4.20 \times 10^{-3}$ | $9.95 \times 10^{-1}$ |
| | Total testosterone | $4.53 \times 10^{-7}$ | $2.92 \times 10^{-6}$ | $3.62 \times 10^{-4}$ | $1.34 \times 10^{-3}$ | $9.98 \times 10^{-1}$ |
| | SHBG | $2.53 \times 10^{-27}$ | $1.63 \times 10^{-26}$ | $1.32 \times 10^{-1}$ | $8.50 \times 10^{-1}$ | $1.80 \times 10^{-2}$ |
| | HDL-c | $2.91 \times 10^{-29}$ | $1.88 \times 10^{-28}$ | $1.32 \times 10^{-1}$ | $8.50 \times 10^{-1}$ | $1.80 \times 10^{-2}$ |
| | LDL-c | $1.02 \times 10^{-48}$ | $6.61 \times 10^{-48}$ | $1.33 \times 10^{-1}$ | $8.58 \times 10^{-1}$ | $8.55 \times 10^{-3}$ |
| | IGF-1 | $1.14 \times 10^{-1}$ | $7.38 \times 10^{-1}$ | $6.48 \times 10^{-4}$ | $4.04 \times 10^{-3}$ | $1.43 \times 10^{-1}$ |
| 2-hour GIP | SHBG (BMI adj.) | $4.52 \times 10^{-41}$ | $6.18 \times 10^{-40}$ | $6.80 \times 10^{-2}$ | $9.31 \times 10^{-1}$ | $1.16 \times 10^{-3}$ |
| | Bioavailable testosterone | $1.12 \times 10^{-6}$ | $1.54 \times 10^{-5}$ | $3.58 \times 10^{-4}$ | $3.91 \times 10^{-3}$ | $9.96 \times 10^{-1}$ |
| | Total testosterone | $2.22 \times 10^{-7}$ | $3.03 \times 10^{-6}$ | $1.77 \times 10^{-4}$ | $1.43 \times 10^{-3}$ | $9.98 \times 10^{-1}$ |
| | SHBG | $1.31 \times 10^{-27}$ | $1.79 \times 10^{-26}$ | $6.80 \times 10^{-2}$ | $9.31 \times 10^{-1}$ | $1.16 \times 10^{-3}$ |
| | HDL-c | $1.50 \times 10^{-29}$ | $2.05 \times 10^{-28}$ | $6.80 \times 10^{-1}$ | $9.31 \times 10^{-1}$ | $1.16 \times 10^{-3}$ |
| | LDL-c | $5.01 \times 10^{-49}$ | $6.85 \times 10^{-48}$ | $6.51 \times 10^{-2}$ | $8.90 \times 10^{-1}$ | $4.50 \times 10^{-2}$ |
| | IGF-1 | $5.76 \times 10^{-2}$ | $7.88 \times 10^{-1}$ | $3.26 \times 10^{-4}$ | $4.31 \times 10^{-3}$ | $1.50 \times 10^{-1}$ |

SHBG = sex hormone-binding globulin, HDL = high density lipoprotein, LDL = low density lipoprotein, IGF-1 = insulin-like growth factor 1
H<sub>0</sub>-H<sub>4</sub>: posterior probabilities of the associations between the 2 traits examined, evaluating 5 different configurations:
- H<sub>0</sub>: Neither trait has an association in the region - H<sub>1</sub>: The first trait has an association in the region but the second does not - H<sub>2</sub>: The second trait has an association in the region but the first does not - H<sub>3</sub>: Both traits have an association in the region but these SNPs are independent - H<sub>4</sub>: Both traits have an association in the region and share a SNP

### Association of traits influenced by E354Q with breast cancer risk

For putative mediators where there was evidence from MR and colocalisation analyses that E354Q influenced that trait, we then evaluated whether there was evidence for an effect of that trait on breast cancer risk. In IVW models, genetically-proxied total testosterone was associated with overall (OR:1.15, 95%CI:1.10-1.21, *P*=9.39×10^−9^), luminal A-like (OR:1.22, 95%CI:1.15-1.30, *P*=5.80×10^−11^), and luminal B HER2 Negative-like breast cancer risk (OR:1.23, 95%CI:1.13-1.34, *P*=1.02×10^−6^).

Likewise, genetically-proxied bioavailable testosterone was associated with overall (OR:1.16, 95%CI:1.04-1.28, *P*=6.53×10^−3^), luminal A-like (OR:1.28, 95% CI:1.14-1.45, *P*=5.27×10^−5^), and luminal B HER2 Negative-like breast cancer risk (OR:1.19, 95%CI:1.14-1.45, *P*=0.02). When employing weighted median and mode models, there was an attenuation of the association of genetically-proxied total testosterone with Luminal B HER2Neg-like breast cancer risk, otherwise findings were broadly consistent across sensitivity analysis models.

We also found evidence that genetically-proxied adult BMI was associated with a lower risk of overall (OR:0.90, 95%CI:0.84-0.96, *P*=1.08×10^−3^), luminal A-like (OR:0.92, 95%CI:0.86-1.00, *P*=0.039), and luminal B HER2Neg-like breast cancer risk (OR:0.89, 95%CI:0.80-0.99, *P*=0.04). Genetically-proxied comparative body size at age 10 was likewise associated with lower risk of overall (OR:0.62, 95%CI:0.55-0.70, *P*=8.25^−14^), luminal A-like (OR:0.65, 95%CI:0.55-0.74, *P*=2.19×10^−8^), and luminal B HER2Neg-like breast cancer risk (OR:0.63, 95%CI:0.52-0.76, *P*=1.87×10^−6^)(**Table 8**). When combining adult BMI and comparative body size at age 10 in a multivariable MR model, the direct effect of adult BMI on breast cancer risk was attenuated for overall and subtype-specific breast cancer (overall breast cancer OR:1.09, 95%CI:0.99-1.20, *P*=0.085) but the direct effect for comparative body size at age 10 was retained for overall and subtype-specific breast cancer (overall breast cancer OR:0.56, 95%CI:0.46-0.67, *P*=5.04×10^−10^)(**Supplementary Table 4**).

**Table 8.** Association between genetically-proxied testosterone (bioavailable and total), glucose levels 2 hours post OGTT, HbA_1c_, T2DM adjusted for BMI, adult BMI, and comparative body size at age 10 and risk of overall and histotype-specific breast cancer.

| Instrument | Outcome | Model | OR (95% CI) | P value |
| --- | --- | --- | --- | --- |
| BT | Breast Cancer Overall | IVW | 1.16 (1.04, 1.28) | 6.53x10<sup>-3</sup> |
|  |  | MR-Egger | 1.16 (0.95, 1.43) | 0.15 |
|  |  | Weighted median | 1.09 (1.01, 1.19) | 0.034 |
|  |  | Weighted mode | 1.10 (1.00, 1.21) | 0.055 |
|  | Luminal A | IVW | 1.28 (1.14, 1.45) | 5.27 x10<sup>-3</sup> |
|  |  | MR-Egger | 1.25 (0.98, 1.58) | 0.072 |
|  |  | Weighted median | 1.24 (1.10, 1.40) | 3.32x10<sup>-4</sup> |
|  |  | Weighted mode | 1.19 (1.04, 1.36) | 0.014 |
|  | Luminal B HER2 Negative | IVW | 1.18 (1.03, 1.36) | 0.020 |
|  |  | MR-Egger | 1.49 (1.14, 1.96) | 4.51 x10<sup>-3</sup> |
|  |  | Weighted median | 1.16 (0.91, 1.47) | 0.24 |
|  |  | Weighted mode | 1.11 (0.85, 1.45) | 0.43 |
| TT | Breast Cancer Overall | IVW | 1.15 (1.10, 1.21) | 9.39 x10<sup>-9</sup> |
|  |  | MR-Egger | 1.17 (1.07, 1.28) | 6.43 x10<sup>-4</sup> |
|  |  | Weighted median | 1.14 (1.07, 1.22) | 2.62 x10<sup>-5</sup> |
|  |  | Weighted mode | 1.15 (1.07, 1.24) | 2.13 x10<sup>-4</sup> |
|  | Luminal A | IVW | 1.22 (1.15, 1.30) | 5.80 x10<sup>-11</sup> |
|  |  | MR-Egger | 1.26 (1.13, 1.41) | 6.04 x10<sup>-5</sup> |
|  |  | Weighted median | 1.21 (1.11, 1.31) | 1.57 x10<sup>-5</sup> |
|  |  | Weighted mode | 1.21 (1.11, 1.33) | 3.63 x10<sup>-5</sup> |
|  | Luminal B HER2 Negative | IVW | 1.23 (1.13, 1.34) | 1.02 x10<sup>-6</sup> |
|  |  | MR-Egger | 1.14 (0.97, 1.33) | 0.10 |
|  |  | Weighted median | 1.25 (1.07, 1.45) | 3.82x10<sup>-3</sup> |
|  |  | Weighted mode | 1.20 (1.03, 1.41) | 0.025 |
| 2hrG | Breast Cancer Overall | IVW | 1.13 (0.97, 1.31) | 0.12 |
|  |  | MR-Egger | 1.41 (0.93, 2.14) | 0.13 |
|  |  | Weighted median | 1.05 (0.94, 1.16) | 0.39 |
|  |  | Weighted mode | 1.03 (0.92, 1.16) | 0.68 |
|  | Luminal A | IVW | 1.10 (0.91, 1.33) | 0.33 |
|  |  | MR-Egger | 1.37 (0.80, 2.37) | 0.28 |
|  |  | Weighted median | 0.98 (0.87, 1.10) | 0.67 |
|  |  | Weighted mode | 0.94 (0.83, 1.08) | 0.41 |
|  | Luminal B HER2 Negative | IVW | 1.18 (1.01, 1.37) | 0.038 |
|  |  | MR-Egger | 1.22 (0.77, 1.91) | 0.41 |
|  |  | Weighted median | 1.09 (0.89, 1.34) | 0.38 |
|  |  | Weighted mode | 1.06 (0.74, 1.50) | 0.77 |
| HbA<sub>1c</sub> | Breast Cancer Overall | IVW | 1.07 (0.87, 1.30) | 0.54 |
|  |  | MR-Egger | 1.01 (0.70, 1.47) | 0.94 |
|  |  | Weighted median | 0.96 (0.81, 1.14) | 0.61 |
|  |  | Weighted mode | 1.00 (0.86, 1.16) | 0.96 |
|  | Luminal A | IVW | 1.04 (0.82, 1.33) | 0.74 |
|  |  | MR-Egger | 0.95 (0.61, 1.49) | 0.84 |
|  | Luminal A | Weighted median | 0.94 (0.75, 1.18) | 0.62 |
|  |  | Weighted mode | 0.94 (0.76, 1.17) | 0.59 |
|  | Luminal B<br>HER2<br>Negative | IVW | 0.96 (0.69, 1.34) | 0.81 |
|  |  | MR-Egger | 0.81 (0.45, 1.48) | 0.50 |
|  |  | Weighted median | 1.05 (0.66, 1.67) | 0.84 |
|  |  | Weighted mode | 0.71 (0.40, 1.26) | 0.25 |
| T2D<br>(BMI adj.) | Breast<br>Cancer<br>Overall | IVW | 0.99 (0.94, 1.05) | 0.78 |
|  |  | MR-Egger | 1.17 (1.04, 1.32) | 0.011 |
|  |  | Weighted median | 1.04 (0.99, 1.09) | 0.13 |
| | | Weighted mode | 1.13 (1.08, 1.20) | $2.32 \times 10^{-5}$ |
|  | Luminal A | IVW | 0.98 (0.91, 1.05) | 0.52 |
| | | MR-Egger | 1.23 (1.08, 1.41) | $3.15 \times 10^{-3}$ |
|  |  | Weighted median | 1.00 (0.93, 1.06) | 0.90 |
| | | Weighted mode | 1.18 (1.09, 1.27) | $6.49 \times 10^{-5}$ |
|  | Luminal B<br>HER2<br>Negative | IVW | 1.01 (0.93, 1.09) | 0.79 |
|  |  | MR-Egger | 1.14 (0.96, 1.35) | 0.15 |
|  |  | Weighted median | 1.09 (0.98, 1.21) | 0.096 |
|  |  | Weighted mode | 1.09 (0.97, 1.21) | 0.16 |
| Adult BMI | Breast<br>Cancer<br>Overall | IVW | 0.90 (0.84, 0.96) | 0.0011 |
| | | MR-Egger | 0.70 (0.59, 0.83) | $3.56 \times 10^{-5}$ |
| | | Weighted median | 0.90 (0.84, 0.96) | $1.82 \times 10^{-3}$ |
|  |  | Weighted mode | 0.95 (0.65, 1.38) | 0.78 |
|  | Luminal A | IVW | 0.92 (0.86, 1.00) | 0.039 |
| | | MR-Egger | 0.74 (0.61, 0.91) | $4.61 \times 10^{-3}$ |
|  |  | Weighted median | 0.97 (0.89, 1.07) | 0.57 |
|  |  | Weighted mode | 1.12 (0.84, 1.49) | 0.46 |
|  | Luminal B<br>HER2<br>Negative | IVW | 0.89 (0.80, 0.99) | 0.040 |
| | | MR-Egger | 0.64 (0.48, 0.85) | $2.23 \times 10^{-3}$ |
|  |  | Weighted median | 0.82 (0.70, 0.95) | 0.010 |
| | | Weighted mode | 0.62 (0.46, 0.84) | $1.96 \times 10^{-3}$ |
| Comparative<br>body size at<br>age 10 | Breast<br>Cancer<br>Overall | IVW | 0.62 (0.55, 0.70) | $8.25 \times 10^{-14}$ |
| | | MR-Egger | 0.49 (0.37, 0.63) | $1.94 \times 10^{-7}$ |
| | | Weighted median | 0.59 (0.52, 0.67) | $2.36 \times 10^{-15}$ |
| | | Weighted mode | 0.55 (0.41, 0.73) | $7.18 \times 10^{-5}$ |
| | Luminal A | IVW | 0.65 (0.55, 0.74) | $2.19 \times 10^{-8}$ |
| | | MR-Egger | 0.51 (0.37, 0.70) | $5.88 \times 10^{-5}$ |
| | | Weighted median | 0.65 (0.56, 0.76) | $4.82 \times 10^{-7}$ |
| | | Weighted mode | 0.62 (0.47, 0.82) | $1.69 \times 10^{-3}$ |
| | Luminal B<br>HER2<br>Negative | IVW | 0.63 (0.52, 0.76) | $1.87 \times 10^{-6}$ |
| | | MR-Egger | 0.47 (0.31, 0.69) | $2.23 \times 10^{-4}$ |
| | | Weighted median | 0.56 (0.42, 0.74) | $4.49 \times 10^{-5}$ |
| | | Weighted mode | 0.52 (0.36, 0.76) | $7.86 \times 10^{-4}$ |

BT = Bioavailable testosterone, TT= Total testosterone, BMI = body mass index, 2hrG= glucose concentration measured 2 hours after OGTT, HbA<sub>1c</sub> = glycated haemoglobin.

There was little evidence for an association of genetically-proxied 2-hour glucose, HbA_1c_ or genetic liability to type 2 diabetes with breast cancer risk (**Table 8**).

## Discussion

In this Mendelian randomization analysis of up to 235,698 cancer cases and 333,932 controls, each copy of the *GIPR* E354Q missense variant was associated with higher risk of overall, luminal A-like, and luminal B HER2 negative-like breast cancer risk. These findings were supported in colocalisation analysis and were replicated in an independent sample of 8,401 breast cancer cases and 99,321 controls. E354Q was also associated with higher 2-hour glucose concentrations but diminished insulin secretion and lower total and bioavailable testosterone concentrations. These measures confer opposing effects on breast cancer risk, suggesting perturbed glycaemic and/or other adverse effects of impaired GIPR signalling through this mechanism offset possible beneficial effects on insulin secretion and circulating testosterone levels. There was little evidence of association of E354Q with risk of the 5 other cancers examined.

The *GIPR* E354Q variant has previously been implicated in increased GIP-GIPR residence time, signalling, internalisation and thus likely desensitisation and downregulation of the signalling pathway long-term in some tissues^23^. Consistent with prior studies, each copy of the E354Q variant was associated with various indices of diminished postprandial insulin secretion^22,56,57^. Given the established role of sustained elevated blood insulin levels in development of breast cancer, the adverse association of E354Q with breast cancer endpoints suggests that this effect is likely mediated via non-insulinemic pathways^9^. This observation is further reinforced by the specificity of the association of E354Q with breast cancer risk, given important roles of hyperinsulinemia in the 5 other cancers examined in this analysis. Though further experimental work is required to validate and clarify potential mechanisms governing this effect, our findings suggesting an adverse association of E354Q with breast cancer risk provide tentative support for a potential protective effect of enhanced GIPR signalling (i.e. GIPR agonism) on breast cancer risk.

Our findings are not consistent with a previous conventional epidemiological analysis which found little evidence of an association of circulating GIP concentrations with breast cancer risk (OR for women at and above vs below median GIP levels: 1.06, 95%CI:0.63-1.84), though this study was restricted to 109 cancer cases and GIP was measured in non-fasting samples which could result in substantial measurement error^17^. While preclinical studies suggest that GIP can induce cAMP elevation in medullary thyroid cancer cells and proliferation in colorectal cancer cells, no known *in vitro* or *in vivo* studies have examined the role of GIP signalling in breast cancer to date^14,58^.

In our analyses, E354Q was associated with lower adult BMI levels which is not consistent with weight loss observed in clinical trials of GIPR agonists (alongside GLP1R agonists)^59^. Interestingly, both GIPR agonists and antagonists have been shown to induce weight loss in preclinical settings^60^. One possible explanation for this apparent paradox is agonism-induced desensitisation of the GIPR, in which persistent stimulation of the GIP receptor by an agonist results in an increasingly diminished response and, consequently, a weight-loss effect^60^. This theory is supported by preclinical work in adipose cell culture which has demonstrated that GIPR responsiveness is impaired following repeated stimulation, and this repeated stimulation results in downregulation of GIPR at the plasma membrane^60,61^.

The E354Q variant was also associated with lower self-reported comparative body size at age 10, but not with measured BMI in children aged 2-10. In univariable MR models, both adult BMI and comparative body size at age 10 were associated with lower breast cancer risk, though only childhood comparative body size showed evidence of a direct effect on breast cancer in multivariable MR models, consistent with prior MR analysis^62^. Consistent with our findings, a recent meta-analysis of 37 prospective studies has also suggested a protective association of higher early life BMI with breast cancer risk^63^. It is therefore plausible that part of a potential adverse effect of E354Q on breast cancer risk is mediated via lower early-life adiposity, though discrepancies in findings between self-reported comparative body size and measured BMI in childhood require further exploration in future studies.

Strengths of this analysis include the use of an Mendelian randomization approach, which should be less susceptible to issues of confounding and reverse causation than conventional epidemiological analyses; the use of a summary-data MR approach which permitted use to leverage data from several large GWAS consortia, increasing statistical power and precision of causal estimates; and the comprehensive assessment of the effect of GIPR signalling across a large panel of glycaemic, hormonal, and lipidomic mediators which enabled us to evaluate potential biological mechanisms through which impaired GIPR signalling may confer an increased risk of breast cancer.

There are several limitations to these analyses. First, drug-target MR analyses are restricted to examining “on-target” effects of pharmacological interventions. Second, effect estimates presented assume linear and time-fixed effects of GIPR signalling and the absence of gene-environment and gene-gene interactions. Third, MR analyses consider the small, lifelong effects exerted by a genetic variant, which may not necessarily translate to the clinical effect observed through pharmacological intervention in adult life. Fourth, statistical power was likely limited for some less common cancer sites (e.g. pancreatic and renal cancer) and histological subtypes (e.g. small cell lung cancer). Statistical power can also often be limited in colocalisation analyses which can reduce the likelihood of shared causal variants across traits being detected. Fifth, we were unable to examine the effect of four measures of insulin secretion (AUC_ins_/AUC_gluc_, AUC_ins_, Ins_30_, and Ins_30_ [BMI adj.]), influenced by E354Q, on breast cancer risk due to the lack of genome-wide significant variants available to serve as instruments for these measures. Sixth, effect estimates were generated from data on participants without type 2 diabetes and therefore findings may not generalise to those with this condition. Finally, while the restriction of participants to those of European ancestry, use of a functional variant in *GIPR* to instrument GIPR signalling, and use of colocalisation should help to minimise exchangeability and exclusion restriction violations, these assumptions are unverifiable.

There is considerable interest in pharmacological modification of GIPR signalling as treatment for type 2 diabetes and obesity. Our findings, using an established missense variant in *GIPR* to proxy impaired GIPR signalling, suggest potential adverse effects of downregulated GIPR signalling on breast cancer risk and, thus, possible protective effects of pharmacological GIPR agonism. Given the sparsity of preclinical and epidemiological literature examining the role of GIPR signalling in breast cancer development, further work is warranted to validate and clarify potential mechanisms underpinning this putative effect. In particular, further evaluation of possible non-insulinemic pathways influenced by GIPR signalling could help to reconcile the specificity of the E354 association with breast cancer risk given the important role of metabolic dysfunction across the 5 other cancers examined in this analysis. Though clinical trial data support the efficacy of dual GIPR/GLP1R agonism for glycaemic control in type 2 diabetes, it is unclear whether pharmacological GIPR agonism alone would confer similar favourable effects on glucose metabolism^59,60^. Evaluation of the role of genetically-proxied GLP1R signalling, alone and in combination with genetically-proxied GIPR signalling, could provide additional insight into the viability of dual pharmacological GLP1R/GIPR agonism for breast cancer prevention.

In conclusion, our drug-target Mendelian randomization analyses across 6 cancers suggest adverse effects of the *GIPR* E354Q missense variant on breast cancer risk. In mechanistic analyses, this variant was associated with higher levels of 2-hour glucose but diminished insulin secretion and lower total and bioavailable testosterone concentrations. Triangulation of these findings in other settings will inform on the efficacy of pharmacologically modifying GIPR signalling as a potential chemoprevention strategy for breast cancer^64^.

## Data Availability

Summary genetic association data for select cancer endpoints were obtained from the public domain: breast cancer (https://bcac.ccge.medschl.cam.ac.uk/bcacdata/), breast cancer in BRCA1/2 mutation carriers (https://cimba.ccge.medschl.cam.ac.uk/projects/), endometrial cancer (https://www.ebi.ac.uk/gwas/), and lung cancer (https://www.ebi.ac.uk/gwas/). Data on pancreatic cancer were obtained via dbGaP release phs000206.v5.p3. Summary genetic association data for colorectal cancer can be accessed by contacting GECCO. Summary genetic association data from the Finngen consortium can be accessed by visiting https://www.finngen.fi/en/access_results. Summary genetic association data from the MAGIC consortium can be obtained by visiting https://magicinvestigators.org/downloads/. Summary genetic association data from the GIANT consortium can be obtained by visitinghttps://portals.broadinstitute.org/collaboration/giant/index.php/GIANT_consortium. Summary genetic association data from the DIAGRAM consortium an be obtained by visiting https://diagram-consortium.org/downloads.html. Summary genetic association data on UK Biobank-derived traits can be accessed via the IEU Open GWAS project (https://gwas.mrcieu.ac.uk/). All other relevant data are within the manuscript and its Supporting Information files.

## Author contributions

Conceptualization: JY. Data Curation: MR, EA, and JY. Formal Analysis: MR. Investigation: all authors. Methodology: MR and JY. Writing—original draft: MR and JY. Writing—review & editing: all authors.

## Acknowledgments

The authors would like to thank the participants of the individual studies contributing to the BCAC, CIMBA, GECCO, CORECT, CCFR, ILCCO, PANC4, MAGIC, DIAGRAM, and GIANT consortia and the UK Biobank and FinnGen study. The authors would also like to acknowledge the investigators of these consortia and studies for generating the data used for this analysis. Summary genetic association data for cancer included data from the following consortia: Breast Cancer Association Consortium (BCAC), Consortium of Investigators of Modifiers of BRCA1/2 (CIMBA), Endometrial Cancer Association Consortium (ECAC), Epidemiology of Endometrial Cancer Consortium (E2C2), UK Biobank, the Genetics and Epidemiology of Colorectal Cancer Consortium (GECCO), the Colorectal Cancer Transdisciplinary Study (CORECT), the Colon Cancer Family Registry (CCFR), the International Lung Cancer Consortium (ILCCO), the Pancreatic Cancer Cohort Consortium (PanScan) and the Pancreatic Cancer Case-Control Consortium (PanC4).

The breast cancer genome-wide association analyses for BCAC and CIMBA were supported by Cancer Research UK (PPRPGM-Nov20\100002, C1287/A10118, C1287/A16563, C1287/A10710, C12292/A20861, C12292/A11174, C1281/A12014, C5047/A8384, C5047/A15007, C5047/A10692, C8197/A16565) and the Gray Foundation, The National Institutes of Health (CA128978, X01HG007492-the DRIVE consortium), the PERSPECTIVE project supported by the Government of Canada through Genome Canada and the Canadian Institutes of Health Research (grant GPH-129344) and the Ministère de l’Économie, Science et Innovation du Québec through Genome Québec and the PSRSIIRI-701 grant, the Quebec Breast Cancer Foundation, the European Community’s Seventh Framework Programme under grant agreement n° 223175 (HEALTH-F2-2009-223175) (COGS), the European Union’s Horizon 2020 Research and Innovation Programme (634935 and 633784), the Post-Cancer GWAS initiative (U19 CA148537, CA148065 and CA148112 - the GAME-ON initiative), the Department of Defence (W81XWH-10-1-0341), the Canadian Institutes of Health Research (CIHR) for the CIHR Team in Familial Risks of Breast Cancer (CRN-87521), the Komen Foundation for the Cure, the Breast Cancer Research Foundation and the Ovarian Cancer Research Fund. All studies and funders are listed in Zhang H et al (Nat Genet, 2020).

The CIMBA data management and data analysis were supported by Cancer Research UK grants C12292/A20861, C12292/A11174. iCOGS: the European Community’s Seventh Framework Programme under grant agreement n° 223175 (HEALTH-F2-2009-223175) (COGS), Cancer Research UK (C1287/A10118, C1287/A 10710, C12292/A11174, C1281/A12014, C5047/A8384, C5047/A15007, C5047/A10692, C8197/A16565), the National Institutes of Health (CA128978) and Post-Cancer GWAS initiative (1U19 CA148537, 1U19 CA148065 and 1U19 CA148112 - the GAME-ON initiative), the Department of Defence (W81XWH-10-1-0341), the Canadian Institutes of Health Research (CIHR) for the CIHR Team in Familial Risks of Breast Cancer (CRN-87521), and the Ministry of Economic Development, Innovation and Export Trade (PSR-SIIRI-701), Komen Foundation for the Cure, the Breast Cancer Research Foundation, and the Ovarian Cancer Research Fund. The PERSPECTIVE project was supported by the Government of Canada through Genome Canada and the Canadian Institutes of Health Research, the Ministry of Economy, Science and Innovation through Genome Québec, and The Quebec Breast Cancer Foundation. All studies and funders are listed in Milne et al (Nat Genet, 2017) and Phelan et al (Nat Genet, 2017).

## Data availability statement

Summary genetic association data for select cancer endpoints were obtained from the public domain: breast cancer (https://bcac.ccge.medschl.cam.ac.uk/bcacdata/), breast cancer in BRCA1/2 mutation carriers (https://cimba.ccge.medschl.cam.ac.uk/projects/), endometrial cancer (https://www.ebi.ac.uk/gwas/), and lung cancer (https://www.ebi.ac.uk/gwas/). Data on pancreatic cancer were obtained via dbGaP release phs000206.v5.p3. Summary genetic association data for colorectal cancer can be accessed by contacting GECCO. Summary genetic association data from the Finngen consortium can be accessed by visiting https://www.finngen.fi/en/access_results. Summary genetic association data from the MAGIC consortium can be obtained by visiting https://magicinvestigators.org/downloads/. Summary genetic association data from the GIANT consortium can be obtained by visiting https://portals.broadinstitute.org/collaboration/giant/index.php/GIANT_consortium. Summary genetic association data from the DIAGRAM consortium an be obtained by visiting https://diagram-consortium.org/downloads.html. Summary genetic association data on UK Biobank-derived traits can be accessed via the IEU Open GWAS project (https://gwas.mrcieu.ac.uk/). All other relevant data are within the manuscript and its Supporting Information files.

## Electronic Resources

PLINK: http://pngu.mgh.harvard.edu/purcell/plink/

LocusZoom: LocusZoom - Create Plots of Genetic Data

